# Quantitative Digitography Solves the Remote Measurement Problem in Parkinson’s disease

**DOI:** 10.1101/2021.11.17.21266483

**Authors:** K.B. Wilkins, M.N. Petrucci, Y. Kehnemouyi, A. Velisar, K. Han, G. Orthlieb, M.H. Trager, J.J. O’Day, S. Aditham, H.M. Bronte-Stewart

## Abstract

**Background:** Assessment of motor signs in Parkinson’s disease (PD) has required an in-person examination. However, 50% of people with PD do not have access to a neurologist. Wearable sensors can provide remote measures of some motor signs but require continuous data acquisition for several days. A major unmet need is reliable metrics of all cardinal motor signs, including rigidity, from a simple short active task that can be performed remotely or in the clinic.

**Objective:** Investigate whether thirty seconds of repetitive alternating finger tapping (RAFT) on a portable quantitative digitography (QDG) device, which measures amplitude and timing, produces reliable metrics of all cardinal motor signs in PD

**Methods:** Ninety-six individuals with PD and forty-two healthy controls performed a thirty-second QDG-RAFT task and clinical motor assessment. Eighteen individuals were followed longitudinally with repeated assessments for an average of three years and up to six years.

**Results:** QDG-RAFT metrics differentiated individuals with PD from controls and provided validated metrics for total motor disability (MDS-UPDRS III) and for rigidity, bradykinesia, tremor, gait impairment and freezing of gait (FOG). Additionally, QDG-RAFT tracked disease progression over several years off therapy, and differentiated akinetic rigid from tremor dominant phenotypes, as well as people with from those without FOG.

**Conclusions:** QDG is a reliable technology, which will improve access to care, allows complex remote disease management, and accurate monitoring of disease progression over time in PD. QDG-RAFT also provides the comprehensive PD motor metrics needed for therapeutic trials.

## Introduction

Parkinson’s disease (PD) is the fastest growing progressive neurological disorder in the world; it has been called an impending pandemic^1^. The treatment of PD requires complex, multi-dose medication regimens and constant monitoring due to adverse effects from medication. The addition of deep brain stimulation (DBS) to the therapy regimen may smooth out some of the medication-induced side effects but also requires frequent visits for DBS programming. The assessment of a person with PD requires an in-person neurological examination to evaluate motor signs of rigidity (stiffness), bradykinesia (slowness), tremor, and gait and balance impairment. Rigidity assessment requires passive range of motion of the patient’s limbs by the examiner and cannot be done via video or with any wearable sensor.

There is no biomarker of PD that can be used to guide therapy decisions. When a person with PD is having difficulty, they must call or message their clinic. Several days may pass before the nurse or physician reaches the person, who then must describe their symptoms in words, which may have changed in the interval. The physician must make complex management decisions based on inadequate information or to try to squeeze in the person in over-booked clinics. Telemedicine has become an established method of care across the world and may offer a solution to this problem. However, assessment of all aspects of motor disability in PD, especially rigidity, is currently impossible via telemedicine^2^. It was noted that without assessment of rigidity there were “no means for direct comparison of remote and in-person MDS-UPDRS III motor scores”^3^. Therefore, a major unmet need is a device that can reliably assess all the cardinal motor signs of PD remotely and transmit validated metrics to the physician in real time.

We originally introduced the technology Quantitative DigitoGraphy (QDG) using the metrics recorded from repetitive alternating finger tapping (RAFT) on a MIDI keyboard and demonstrated that it was a validated objective measure of overall motor impairment and motor signs in PD; it differentiated people with PD from controls, PD off and on therapy, and reflected tremor and freezing events^4,5^. The MIDI keyboard primarily registered only full amplitude strikes. Although it reported a proprietary measure of the force of strike, the validated metrics were otherwise in the temporal domain, such as the frequency of tapping, keystrike duration, and the variability in keystrike duration and frequency (arrhythmicity), similar to finger tapping tasks implemented on smart phones^4–6^. We subsequently developed an engineered digitography device, that can accurately track amplitude displacement and temporal metrics during the task since amplitude is a critical component of PD-related movement^7^. We demonstrated that keystrike amplitude and amplitude variability reliably distinguished people with moderate PD and people with chronic HIV from controls, and the slope of the release amplitude was a robust measure of rigidity in PD^8,9^.

This study, in a large cohort of people with PD and healthy controls, demonstrates that thirty seconds of RAFT on an engineered digitography device reliably differentiates these groups, provides validated metrics of all the major cardinal motor signs of PD, including rigidity, bradykinesia, tremor, gait impairment, and freezing of gait (FOG), and tracks progression of such metrics over time. Additionally, this study shows that QDG-RAFT metrics can distinguish the akinetic rigid from the tremor dominant PD phenotypes, and people with from those without FOG. A simple, short QDG-RAFT task, analyzed by an automated algorithm in real time, will improve access to care and will provide reliable comprehensive data to enable complex disease management decisions via telemedicine. It will enable remote monitoring of disease progression over time, without requiring additional in person visits and will provide objective metrics of all motor signs including rigidity, which solves a critical unmet need in video-based outcomes in clinical trials. QDG technology provides a unique and valuable companion to the suite of wearable sensors that are only validated for a subset of motor symptoms and require continuous data acquisition over the course of hours to days.

## Materials and Methods

### Human Subjects

Ninety-six individuals (52 males, 44 females) with clinically established Parkinson’s disease^10^ and forty-two healthy controls (20 males, 22 females) participated in the study. All experimental testing was done in the off-therapy state. Long- and short-acting medications were withdrawn over 24 to 48 and 12 h, respectively. If the patient had DBS, therapy was turned off for at least 15 min before testing began^11^. All participants gave written informed consent to participate in the study, which was approved by the Stanford University Institutional Review Board.

### Experimental Protocol

Individuals performed repetitive alternating finger tapping (RAFT) on tensioned, engineered keys on a digitography device, which senses the amplitude displacement and timing of keystrikes. Individuals sat with their wrist resting on a pad at the same level as the keys of the device. They placed the index and middle finger on adjacent keys. The instructions were to press and release each key in an alternating pattern, as fast and regularly as possible for thirty seconds, starting and stopping only when they heard an auditory cue. They were instructed to attempt to press and release the keys completely. Participants performed the tasks with their eyes closed and while listening to white noise over headphones to remove visual and auditory feedback. Each subject had a short period of practice before the test began. Patients performed the task with each hand.

In addition to the RAFT task, individuals also performed the motor scale of the Unified Parkinson’s Disease Rating Scale (UPDRS III). Thirteen individuals performed the original UPDRS III and 83 individuals performed the Movement Disorders Society-UPDRS III (MDS-UPDRS III). Sub-scores for the UPDRS were defined as follows: 1. Bradykinesia: sum of finger tapping, hand movements, and pronation/supination items for the tested upper extremity, 2. Rigidity: rigidity item for the tested upper extremity, 3. Tremor: sum of postural and resting tremor items of the tested arm, 4. Gait & Freezing of Gait (FOG): sum of gait and freezing of gait items. An individual was considered a freezer if they scored >0 on either the Freezing of Gait Questionnaire (FOGQ) or exhibited freezing during gait assessments in the lab.

A subset of this cohort (N=18) was followed longitudinally with multiple performances of RAFT to assess changes in each of the QDG metrics over time. Tests were separated by a minimum of 3 months.

### Kinematic Data Acquisition and Analysis

The device produced a voltage signal that was proportional to the displacement of the key. The key displacement was linearly related to the output voltage signal with a resolution of 62.5 μm per 40 mV. For key displacements less than 9 mm, the device operated in a linear zone. Near the base of the key displacement, the key reached a compliant mechanical stop where displacement was non-linearly related to the output voltage signal. A customized detection algorithm was used to determine specific states in the cycle of finger movement (Figure 1A). Six metrics were then calculated from these cycles: 1. Press amplitude, 2. Press amplitude coefficient of variation (CV: standard deviation/mean), 3. Inter-strike interval (ISI: time to complete one cycle of finger movement), 4. Inter-strike interval CV (ISI CV), 5. Release slope (i.e., ratio of the amplitude of the key release compared to duration of release), and 6. Dwell time (i.e., duration at bottom of the press). Release amplitude and release amplitude CV were not investigated due to high collinearity with press amplitude (R>0.99) and press amplitude CV (R>0.99). The average for each of these metrics across the thirty second trial was computed for each finger and then averaged across fingers for each hand.

**Figure 1.**
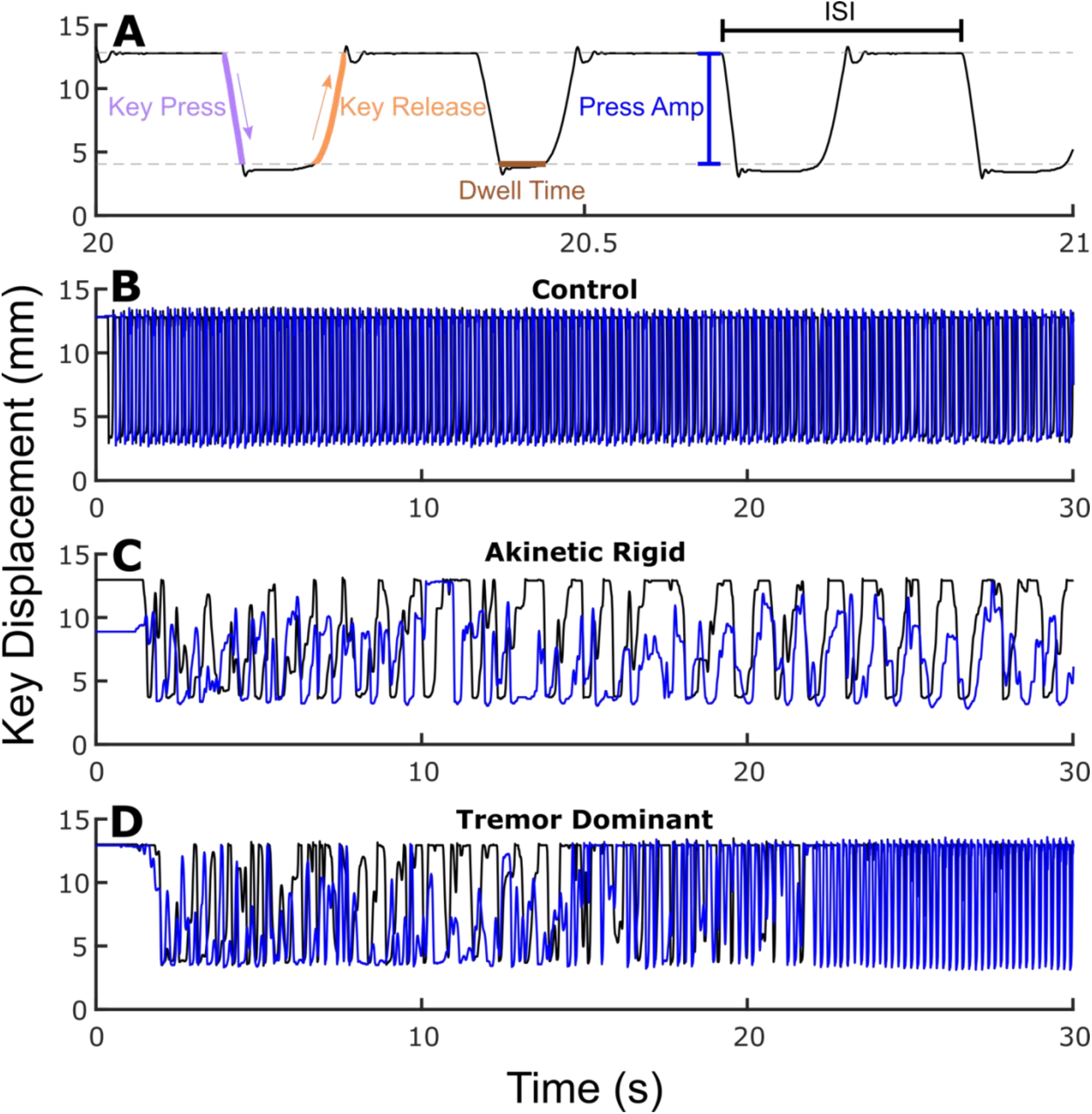
(A) Zoomed in QDG trace. Gray dashed lines represent nonlinear zones. Examples of (B) healthy control, (C) akinetic rigid PD phenotype, and (D) tremor dominant PD phenotype with one finger in blue and the other in black.

### Statistical Analysis

Statistical analyses were run in R (version 3.6.0, R Foundation for Statistical Computing, University of Auckland, New Zealand) and MATLAB (version 9.9, Mathworks, Natick MA). To assess differences in the six metrics between individuals with PD and healthy controls, a linear mixed effects model was used with fixed effects of group (PD vs. C) and age, as well as a random intercept for subject. A Pearson correlation was computed between total MDS-UPDRS III and each of the 6 metrics. Spearman correlations were used to assess correlations between sub-scores of the UPDRS and the 6 metrics. To assess changes over time, a linear mixed effects model was used with fixed effects of time and age, a random slope of time, and random intercept for subject. To measure differences in phenotypes (i.e., akinetic rigid [AR] vs. tremor dominant [TD]), a linear mixed effects model was performed on the longitudinal cohort with a fixed effect of phenotype and age, along with a random slope of time and random intercept for subject for each of the 6 metrics. Lastly, a similar linear mixed effects model but with a fixed effect of subtype (freezer vs. non-freezer) was calculated to evaluate differences for each of the 6 metrics between freezers and non-freezers. Significance was set at *p* < 0.05. All reported p-values were corrected with a Bonferroni correction for the six metrics tested.

## Results

### Demographic Characteristics

Demographic information for the individuals with PD and Controls is shown in Table 1. The PD group had a mean MDS-UPDRS III (N=83) of 25.1 ± 13.4 and disease duration of 8.7 ± 5.7 years. The healthy controls were younger (60.0 ± 9.0 years) on average than the participants with PD (65.1 ± 9.1 years), but age was controlled for when evaluating group differences in metrics.

**Table 1.** Participant Demographics

|  | **Controls** | **PD** |
| --- | --- | --- |
| N | 42 | 96 |
| Age | 60.0 ± 9.0 [44.0 - 76.0] | 65.1 ± 9.1 [34.4 - 90] |
| Sex | 20 M, 22 F | 52 M, 44 F |
| MDS-UPDRS III | - | 25.1 ± 13.4 [4 - 71] |
| UPDRS III | - | 31.6 ± 9.7 [12 - 49] |
| Disease Duration | - | 8.7 ± 5.7 [1 - 22] |

### Finger Tapping Metrics Differ Between Individuals with Parkinson’s disease and Healthy Controls

Figure 1 depicts individual QDG-RAFT traces from a healthy control (Fig 1B) and two individuals with PD, one with the akinetic rigid (AR) phenotype (Fig 1C) and one with the tremor dominant (TD) phenotype (Fig 1D).

The alternating finger tapping in the healthy control is characterized by rhythmic, rapid, full amplitude keystrikes for the duration of the thirty seconds. The AR individual displays small and variable amplitudes of presses and releases throughout the trace, slower overall tapping speed (longer ISIs), and a slower release slope of the key. However, they maintain an alternating finger tapping pattern. The TD individual, Figure 1D, also initially displays alternating finger tapping, with variability in tapping amplitude. After ∼20 seconds, involuntary tremor over-rode the voluntary tapping as evident by the high frequency (>4 Hz), full amplitude, short duration, non-alternating strikes.

Group differences between PD and controls for each of the six metrics were explored, while controlling for age, Figure 2. Press amplitude (t = 6.60, *p* = 1.62e-09), press amplitude CV (t = 7.96, *p* = 4.53e-13), ISI CV (t = 7.94, *p* = 5.54e-13), release slope (t = 7.20, *p* = 4.97e-11), and ISI (t = 2.83, *p* = 0.030) showed significant differences between PD and controls, Figure 1A-E. There was no difference in Dwell time (t = 1.49, *p* = 0.83) between groups, Figure 1F. An increase in the coefficient of variation (CV) in a metric signifies greater variability.

**Figure 2.**
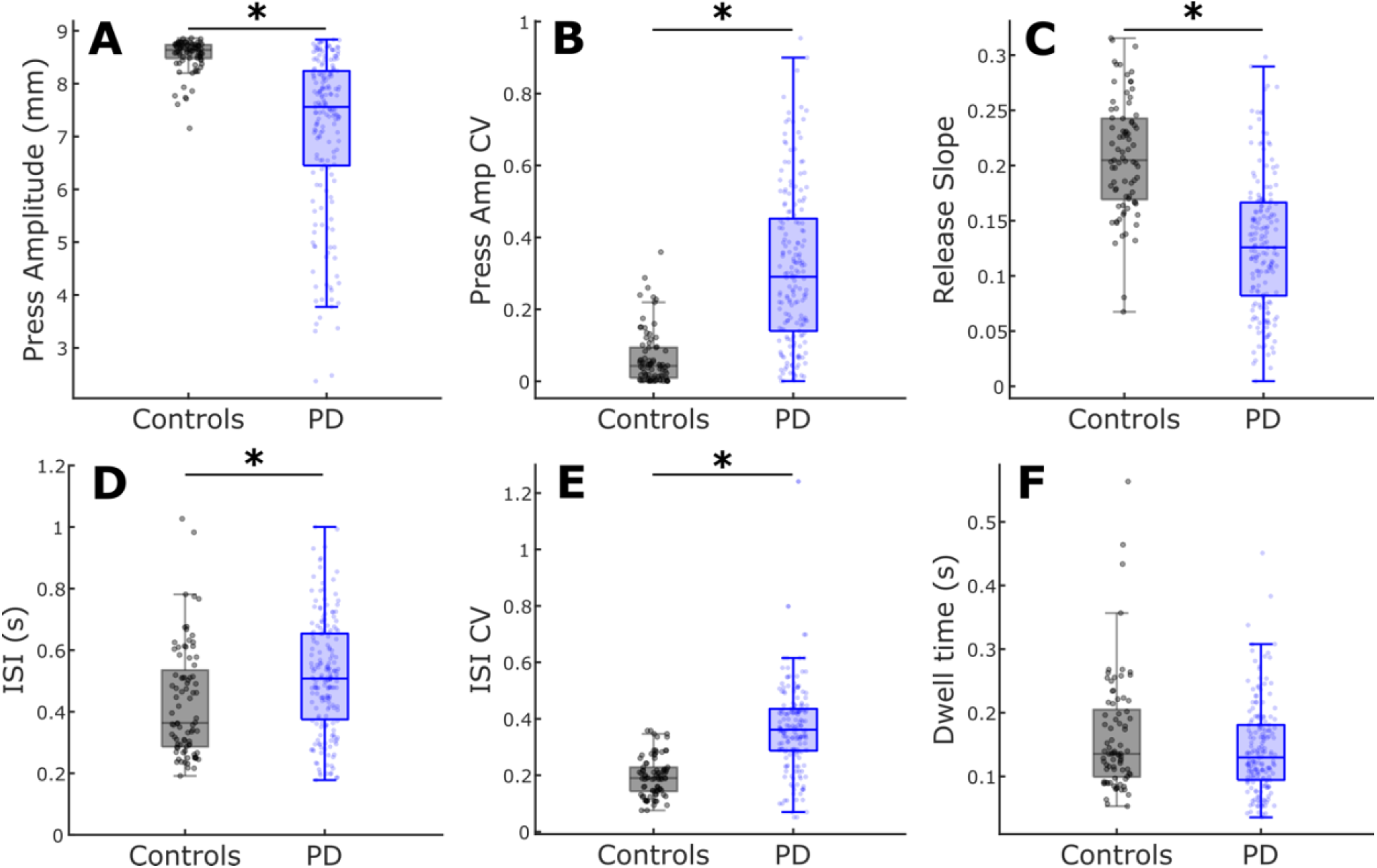
Comparison of QDG metrics between PD and controls. Boxplots with individual data overlaid for (A) Press amplitude, (B) Press amplitude CV, (C) Release slope, (D) ISI, (E) ISI CV, and (F) Dwell time. *indicates significant differences between groups.

### QDG-RAFT produces validated metrics of cardinal motor signs

Individuals who performed the original UPDRS (N=26) were not included in the correlations with the total MDS-UPDRS III or gait & FOG score. There were significant associations between total MDS-UPDRS III (N=166) and press amplitude (R = -0.38, *p* = 5.71e-6), ISI CV (R = 0.36, *p* = 3.50e-5), press amplitude CV (R = 0.26, *p* = 7.20e-3), and release slope (R = -0.23, *p* = 0.029). Dwell time and ISI did not show a significant association, Table 2.

**Table 2.** Correlation Results

| UPDRS Score | QDG Metric | Correlation Coef | p-Value (Corrected) |
| --- | --- | --- | --- |
| Total | **Press Amplitude** | **-0.38** | **5.71e-6** |
|  | **ISI CV** | **0.36** | **3.50e-5** |
|  | **Press Amplitude CV** | **0.26** | **7.20e-3** |
|  | **Release Slope** | **-0.23** | **0.029** |
|  | ISI | 0.19 | 0.076 |
|  | Dwell Time | 0.19 | 0.13 |
| Rigidity<br>Sub-score | **Release Slope** | **-0.43** | **9.78e-9** |
|  | **Dwell Time** | **0.27** | **1.13e-3** |
|  | **Press Amplitude** | **-0.25** | **4.23e-3** |
|  | **ISI** | **0.21** | **0.021** |
|  | **Press Amplitude CV** | **0.21** | **0.029** |
|  | ISI CV | -0.070 | 1.00 |
| Bradykinesia<br>Sub-score | **Press Amplitude** | **-0.34** | **1.56e-5** |
|  | **Release Slope** | **-0.33** | **2.47e-5** |
|  | **Press Amplitude CV** | **0.28** | **7.08e-4** |
|  | ISI CV | 0.15 | 0.31 |
|  | Dwell Time | 0.12 | 0.72 |
|  | ISI | -0.018 | 1.00 |
| Tremor<br>Sub-score | **Dwell Time** | **-0.26** | **2.94e-3** |
|  | ISI | -0.13 | 0.43 |
|  | Release Slope | 0.13 | 0.48 |
|  | ISI CV | 0.10 | 1.00 |
|  | Press Amplitude CV | 0.10 | 1.00 |
|  | Press Amplitude | -0.069 | 1.00 |
| Gait & FOG<br>Sub-score | **ISI CV** | **0.28** | **2.86e-3** |
|  | **Release Slope** | **-0.26** | **7.80e-3** |
|  | Press Amplitude | 0.21 | 0.050 |
|  | Press Amplitude CV | 0.16 | 0.31 |
|  | Dwell Time | 0.14 | 0.45 |
|  | ISI | 0.055 | 1.00 |

To determine if there was any relationship between QDG-RAFT metrics and specific cardinal motor signs, we quantified correlations between rigidity, bradykinesia, and tremor sub-scores across both hands of the full cohort (N = 192), Table 2. There were significant associations between the rigidity sub-score and release slope (ρ = -0.43, *p* = 9.78e-9), Dwell time (ρ = 0.27, *p* = 1.13e-3), press amplitude (ρ = -0.25, *p* = 4.23e-3), ISI (ρ = 0.21, *p* = 0.021), and press amplitude CV (ρ = 0.21, *p* = 0.029). The bradykinesia sub-score significantly correlated with press amplitude (ρ = -0.34, *p* = 1.56e-5), release slope (ρ = -0.33, *p* = 2.47e-5), and press amplitude CV (ρ = 0.28, *p* = 7.08e-4). Tremor sub-score was significantly associated with Dwell time (ρ = -0.26, *p* = 2.94e-3). Gait & FOG sub-scores (MDS-UPDRS III cohort only) significantly correlated with ISI CV (ρ = 0.28, *p* = 2.86-3) and release slope (ρ = -0.26, *p* = 7.80-3).

### QDG-RAFT Performance Deteriorates Over Time

Eighteen individuals were followed longitudinally to assess changes in QDG metrics over time, Figure 3. This group was followed for an average of 32.9 ± 26.2 months, and up to six years, with an average number of visits over that time of 4.5 ± 2.6. Press amplitude (t = 4.02, *p* = 5.36e-4), press amplitude CV (t = 3.91, *p* = 8.30e-4), release slope (t = 4.77, *p* = 2.49e-5), ISI (t = 3.96, *p* = 6.86e-4), and ISI CV (t = 3.78, *p* = 1.35e-3) all significantly worsened over time, Figure 3. Meanwhile, Dwell time (t = 1.78, *p* = 0.46) did not significantly change.

**Figure 3.**
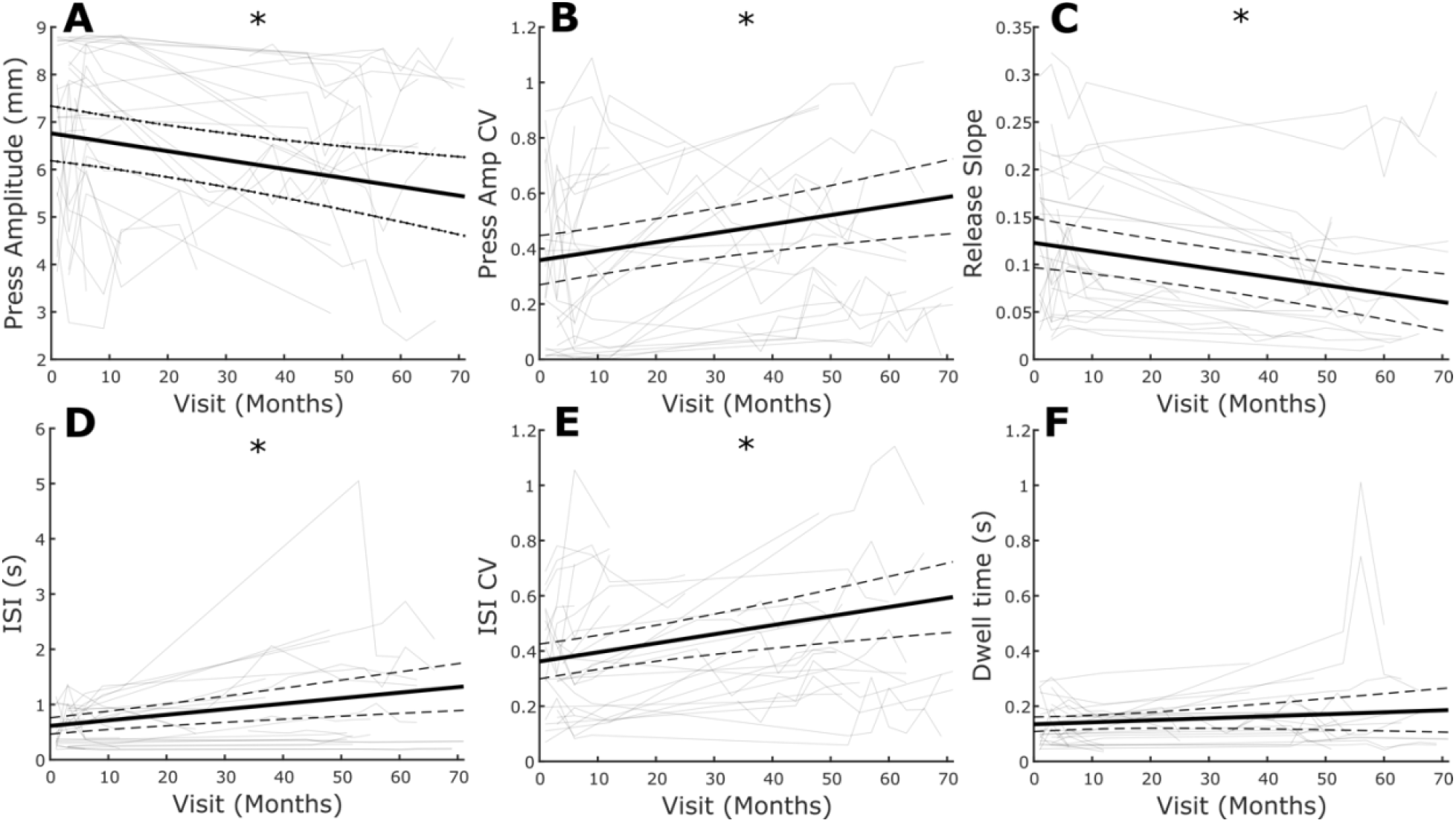
Average change over time (thick black line) with individual data overlaid (light gray) for (A) Press amplitude, (B) Press amplitude CV, (C) Release slope, (D) ISI, (E) ISI CV, and (F) Dwell time. *indicates significance. Dashed lines represent 95% confidence interval.

### QDG-RAFT metrics distinguished between AR and TD PD phenotypes

Within the longitudinal cohort, there was a significant difference between AR and TD phenotypes for release slope (t = 2.74, *p* = 0.041) and Dwell time (t = 3.41, *p* = 5.03e-3), Figure 4. The AR phenotype demonstrated slower release slopes and longer keystrike durations that the TD phenotypes. There were no differences between phenotypes for press amplitude, press amplitude CV, ISI, or ISI CV. These results remained consistent when controlling for total motor impairment by including total MDS-UPDRS III as an additional fixed effect in the mixed model (see Supplementary Table 1).

**Figure 4.**
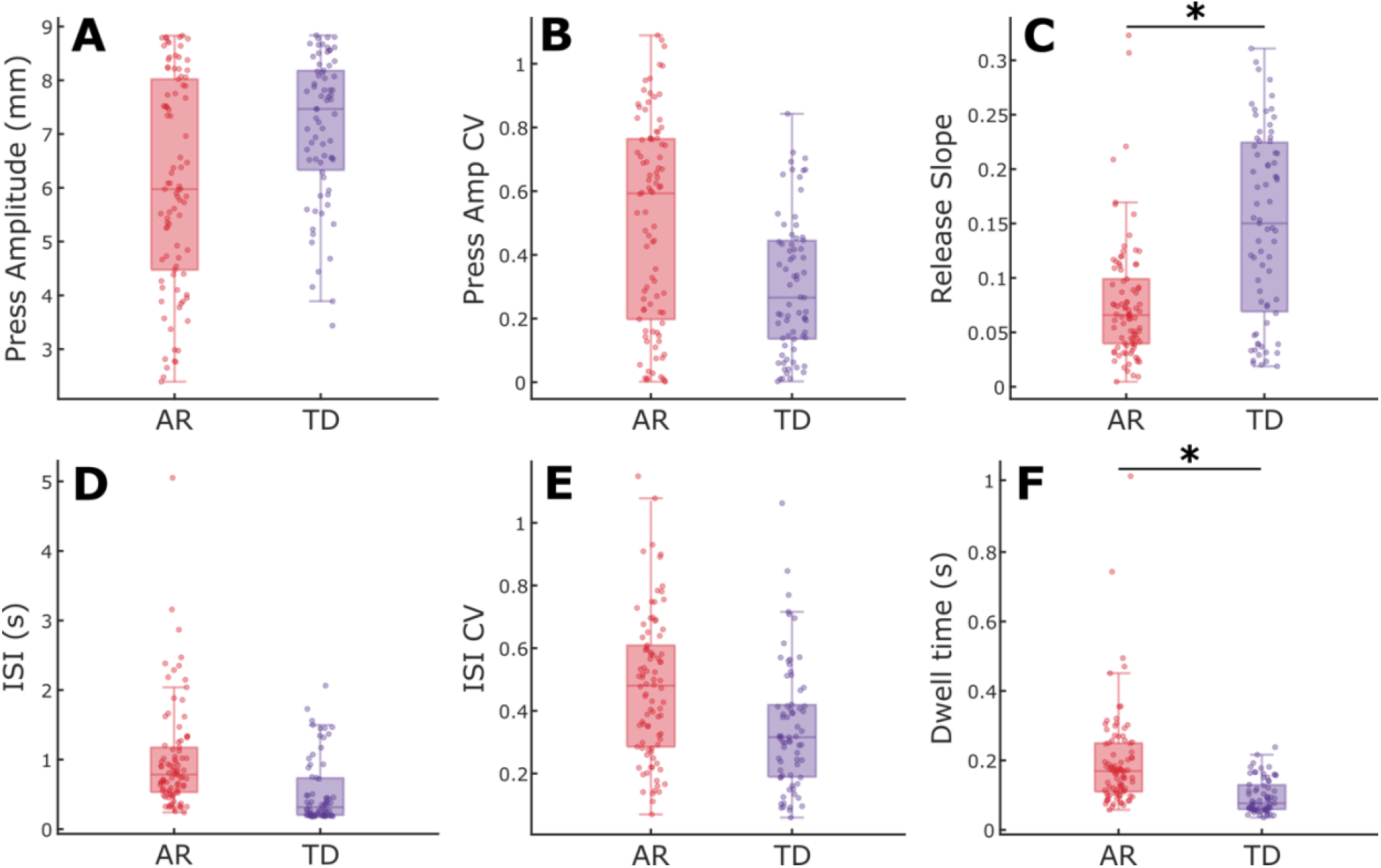
Comparison of QDG metrics between akinetic rigid and tremor dominant phenotypes. Boxplots with individual data overlaid for (A) Press amplitude, (B) Press amplitude CV, (C) Release slope, (D) ISI, (E) ISI CV, and (F) Dwell time. *indicates significant differences between groups.

### Freezers Demonstrate Worse Finger Tapping Performance Compared to Non-Freezers

A subset of the longitudinal cohort (N=15) was used to assess differences between individuals with FOG (freezers) and those without evidence of FOG (non-freezers), Figure 5. Individuals who converted from a non-freezer to a freezer during the period (N=3) were excluded from the analysis. Freezers were significantly worse across all six QDG metrics, including press amplitude (t = 3.22, *p* = 0.010), press amplitude CV (t = 3.47, *p* = 4.37e-3), release slope (t = 4.32, *p* =1.90e-4), ISI (t = 3.89, *p* = 9.82e-4), ISI CV (t = 2.90, *p* = 0.026), and Dwell time (t = 4.55, *p* = 7.80e-5). These results remained consistent when controlling for total motor impairment by including total MDS-UPDRS III as an additional fixed effect in the mixed model (see Supplementary Table 2).

**Figure 5.**
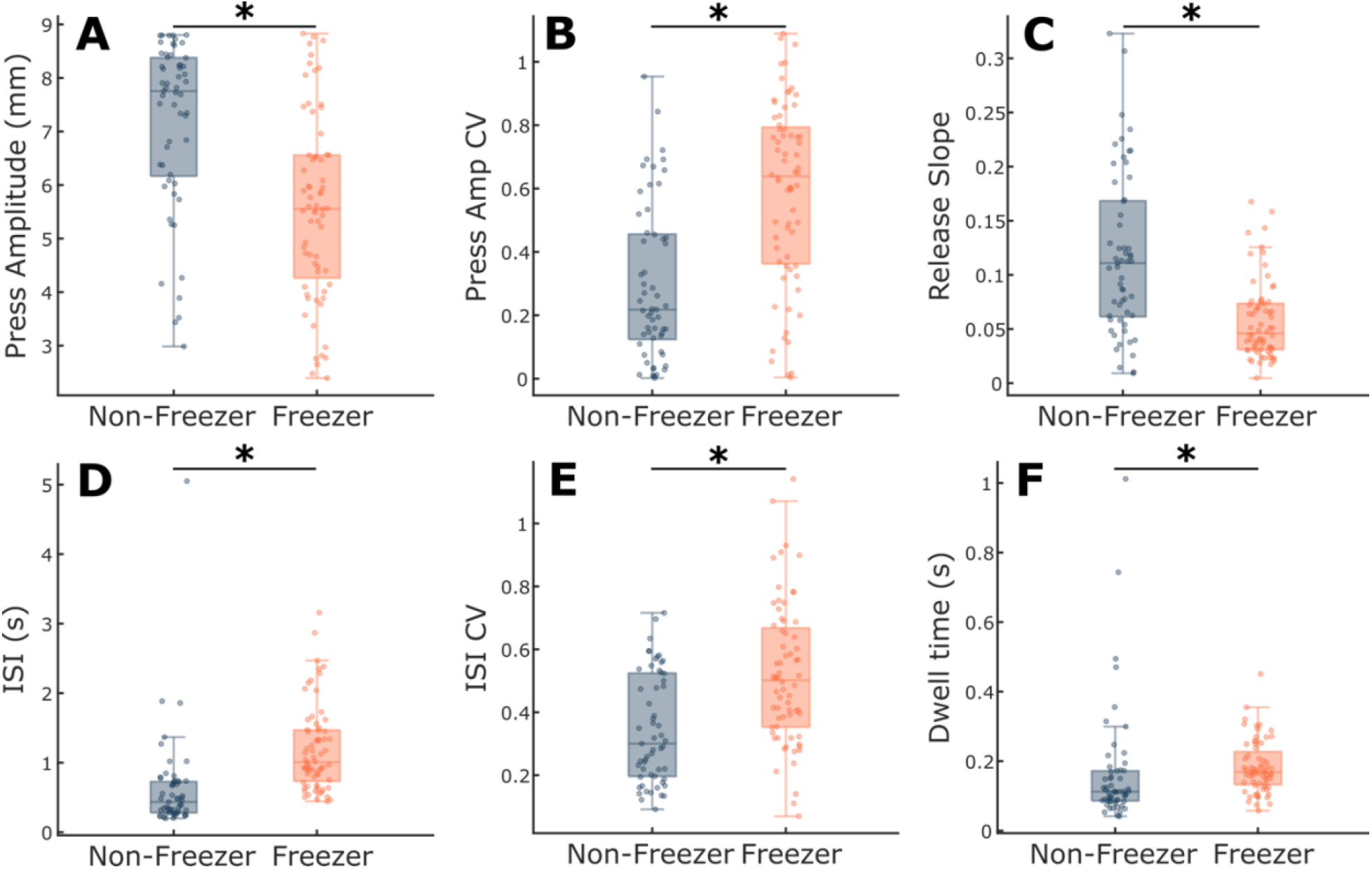
Comparison of QDG metrics between freezer and non-freezer subtypes. Boxplots with individual data overlaid for (A) Press amplitude, (B) Press amplitude CV, (C) Release slope, (D) ISI, (E) ISI CV, and (F) Dwell time. *indicates significant differences between groups.

## Discussion

The current large study has demonstrated that a thirty second repetitive alternating finger tapping (RAFT) task on an engineered digitography device that accurately measures amplitude and timing reliably differentiated people with Parkinson’s disease (PD) from healthy controls. Quantitative digitography (QDG) produced metrics that were validated with overall motor disability (total MDS-UPDRS III score), and with all the major cardinal motor signs of Parkinson’s disease, including rigidity, bradykinesia, tremor, gait impairment and FOG. QDG technology reliably tracked motor progression over several years, off therapy. Furthermore, QDG-RAFT metrics differentiated between akinetic rigid and tremor dominant phenotypes, as well as between freezers and non-freezers. Together, these results validate the use of portable QDG-RAFT as a remote, reliable, and comprehensive tool, which can improve access to care for people with PD and provide quantitative validated data for complex medical management decisions. The task is simple, takes thirty seconds, and the analyzed data is provided in real time. QDG technology and the rigidity metric provide a critically important supplement to video MDS-UPDRS assessments of PD in clinical trials.

### Differentiating PD from Healthy Controls

Individuals with PD had significantly smaller and more variable press amplitudes, slower release amplitude slopes, and slower and more arrhythmic tapping than age matched healthy controls. These findings build on prior work showing the effectiveness of an alternating finger tapping task to differentiate individuals with PD from controls^4,5,9,12^. In fact, alternating finger tapping has been shown to be the most effective motor predictor for conversion from idiopathic REM sleep behavior disorder to Parkinson’s disease, with deficits observed nine years prior to conversion^13^. The effectiveness of differentiating PD from controls has led to the widespread implementation of finger tapping on smart phone battery of assessments for PD^12,14–17^. However, smartphone applications are limited to temporal metrics of tapping and position in 2D space of the fingers^15^. Our current results demonstrate that amplitude-based metrics are critical components for differentiating aspects of Parkinsonism motor deficits, especially rigidity and bradykinesia.

### QDG-RAFT validation to the MDS-UPDRS III and its sub-scores

One of the major results from this study is that the 30 second QDG-RAFT was able to generate metrics that significantly correlated with overall motor impairment in PD, as well as with the major cardinal signs of PD: rigidity, bradykinesia, tremor, gait impairment and FOG. Our lab recently demonstrated that the QDG-RAFT release slope significantly correlated with rigidity of the upper extremity in a smaller cohort^9^. One possible mechanism is that the release of the key is partly a passive movement: the tensioned key partially pushes the finger up as the key is restored to its neutral position. Increased rigidity or stiffness will lead to slower releases and lower release slopes. The ability to quantitatively assess rigidity is a major advancement since rigidity cannot be assessed remotely via video. Successful efforts to develop remote devices for assessing rigidity have been limited and none can also measure the other cardinal PD motor signs^18–22^. Typical wearable inertial monitoring units (IMUs) or smart watches use spectral and kinematic analyses of movement and are unable to measure rigidity^14,23^.

Lower press amplitude and variability in both the amplitude and temporal domains were critical metrics related to the MDS-UPDRS bradykinesia score, which represents the well known hypometria and sequence effect in clinical assessments of repetitive finger tapping and other movements in PD. In QDG these can now be assessed independently unlike in the MDS-UPDRS III.

Higher tremor scores were related to shorter Dwell times during RAFT. This is a result of the involuntary nature of tremor, which when manifest on the keys, produces rhythmic involuntary high frequency tapping with a very short period of time between the press and release phases (Dwell time).

QDG-RAFT arrhythmicity and release slope were significantly correlated with the MDS-UPDRS III items of gait impairment and FOG. Arrhythmicity of gait is a fundamental aspect of gait impairment and a predictor of FOG, falls and related postural deficits^24,25^. This demonstrates the system-wide nature of aberrant motor control in PD: metrics from simple RAFT task are markers of a critical aspect of axial motor disability and may be used as surrogates in remote assessments and monitoring.

### QDG-RAFT documents the progression of PD motor disability over several years

To our knowledge, this is the first study of its kind to track quantitative metrics related to the major cardinal signs of PD longitudinally for several years. We found that individuals got slower, more irregular, stiffer, and moved with smaller amplitude, as would be expected with progressive parkinsonism in the off-therapy state. Although longitudinal quantitative tracking has been successfully applied to aspects of gait^28–30^, assessments of overall motor impairment and symptoms have been mainly limited to clinical assessments that have low reliability^31^. The MM4PD smart watch system was used to track symptoms over time, but that was limited to six months and only examined tremor and dyskinesias on therapy^14^. Analysis of the hold and release times of daily computer keyboard use in people with PD was evaluated to document treatment response over time, but the keyboard metric was not related to any PD motor sign and the duration of the longitudinal study was only six months^32^. To date, most studies investigating disease progression over time have used the MDS-UPDRS III^33^. While this is a comprehensive clinical assessment, the substantial intra- and inter-rater variability and the requirement for an in-person evaluation demonstrates the need for an additional rapid, remote, and reliable technology that tracks progression of all cardinal motor signs^31^. Incorporating QDG technology into clinical practice and clinical trials marks a valuable step forward in being able to both understand progression of different Parkinsonian symptoms over time and create data-informed personalized treatment plans for optimizing therapy.

### QDG-RAFT differentiates between the akinetic-rigid and tremor-dominant phenotypes and can identify people with Freezing of Gait

QDG-RAFT was also able to differentiate between akinetic rigid (AR) and tremor dominant (TD) PD phenotypes. Specifically, the AR cohort had slower release slopes, indicative of greater rigidity, and the TD cohort had shorter Dwell time, suggesting greater number of short duration strikes. These findings matched the initial correlations between QDG metrics and UPDRS sub-scores. We originally identified short durations of keystrikes on a MIDI keyboard, along with tapping frequencies of 4+ Hz as signatures of tremor^4^. Interestingly lower release slopes were also correlated with gait impairment and FOG and AR phenotypes may be more likely to exhibit gait deficits^26,27^.

PD participants identified as freezers from gait tasks and the FOGQ were significantly worse in their performance of QDG-RAFT amplitude, release slope, frequency, and regularity metrics than non-freezers, even when controlling for overall motor impairment. The topic of whether freezing of upper limb (FO-UL) movement is related to FOG has been debated^34–38^. Arrhythmicity of gait (stride time CV) is a robust correlate of FOG^39–41^. A study of bimanual repetitive drawing did demonstrate episodes of FO-UL correlated with FOG, but without major arrhythmicity^34^. We believe a complex unimanual task may be more effective for evoking freezing behavior since we previously found that unimanual QDG, but not bimanual single finger tapping, elicited arrhthmicity^4,7^. The significance of all six QDG-RAFT metrics likely reflects the different aspects of freezing: e.g., cessation of movement (reflected in longer ISI and smaller amplitude of presses) and arrhythmicity (reflected in higher press amplitude CV and ISI CV).

### Limitations

The testing here was limited to off-therapy and therefore dyskinesias were not present during any of the testing. Future on-therapy studies could assess whether it is possible to assess dyskinesia in the tested limb with QDG metrics.

## Conclusion

A simple, short QDG-RAFT task, analyzed by an automated algorithm in real time, provided quantitative metrics of all the cardinal motor signs of PD, including rigidity and gait impairment/FOG, tracked disease progression over time, and differentiated among PD phenotypes. Remote use of QDG technology will improve access to care and complex disease management decisions via telemedicine. This will allow fewer in-person visits without compromising quality of care. It helps solve the critical unmet need for remote comprehensive assessment of PD motor symptoms to inform treatment plans that are currently unaddressed by available wearables.

## Data Availability

All data produced in the present study are available upon reasonable request to the authors

## Acknowledgements

We would like to thank the members of the Human Motor Control and Neuromodulation lab, especially Shannon Hoffman and Hannah Dorris for project feedback, and, most importantly, the participants who dedicated their time to this study.

