## Supplemental Materials for "Quantitative Digitography Solves the Remote Measurement Problem in Parkinson’s disease"

Supplementary Table 1. Phenotype Differences Corrected for Impairment Level

| **QDG Metric** | **T-Value** | **p-Value (Corrected)** |
| --- | --- | --- |
| **Dwell Time** | **4.17** | **2.96e-4** |
| **Release Speed** | **2.80** | **0.034** |
| Press Amplitude CV | 2.08 | 0.24 |
| ISI | 1.94 | 0.33 |
| Press Amplitude | 1.86 | 0.39 |
| ISI CV | 1.32 | 1.00 |

Supplementary Table 2. Freezer Differences Corrected for Impairment Level

| **QDG Metric** | **T-Value** | **p-Value (Corrected)** |
| --- | --- | --- |
| **Release Speed** | **4.34** | **1.80e-4** |
| **Dwell Time** | **3.87** | **0.0010** |
| **Press Amplitude CV** | **3.08** | **0.015** |
| **ISI** | **2.96** | **0.023** |
| **ISI CV** | **2.91** | **0.026** |
| **Press Amplitude** | **2.69** | **0.049** |
